# Burden of febrile illness complications and determinants of typhoid fever complications in the Ashanti region of Ghana

**DOI:** 10.1101/2025.04.17.25326003

**Authors:** Portia Boakye Okyere, Sampson Twumasi-Ankrah, Sam Newton, Samuel Nkansah Darko, Kennedy Gyau Boahen, Michael Owusu, Yeonseon Joanne Kim, Hyon Jin Jeon, Florian Marks, Yaw Adu-Sarkodie, Ellis Owusu-Dabo

## Abstract

**Background and Aim:** Typhoid fever remains a significant global public health concern and ranked among the top ten causes of outpatient illnesses in Ghana from 2020 to 2022. Identifying the risk factors associated with its complications is crucial for reducing mortality. This study aimed to assess the prevalence, types, and risk factors of complications in febrile patients, with a specific focus on typhoid fever in the Ashanti region of Ghana.

**Method:** Clinical data from 238 patients with positive blood cultures, obtained from the surveillance for Severe Typhoid in Africa program between May 2016 and September 2019 were analysed. Statistical reduction techniques were used to identify key variables, and logistic regression identified factors independently associated with typhoid complications.

**Results:** The overall prevalence of complications amongst the febrile patients was 55% (131/238). *S.* Typhi and non-*S.* Typhi infections resulted in 38.9% (28/72) and 62% (103/166) complications, respectively. In general, notable complications associated with febrile illnesses included sepsis (23.5%, 56/238), severe anaemia (19.3%, 46/238), gastrointestinal bleeding (3.8%, 9/238), and intestinal perforation (2.5%, 6/238). Typhoid fever contributed to 18.1% (13/72) of sepsis, 8.3% (6/72) of gastrointestinal bleeding, and 5.6% (4/72) each for severe anaemia and intestinal perforation. Mortality rates were 1.4% (1/72) in *S.* Typhi patients and 2.4% (4/166) in non-*S.* Typhi patients. Complications were independently associated with a longer duration of illness before hospital visit (AOR 8.6, 95%CI 1.2-64.7, p=0.035) and rural residence (AOR 0.1, 95%CI 0, 0.7, p=0.024).

**Conclusion:** The study highlights a significant burden of complications among febrile patients in Ghana, with typhoid fever as a major concern. Delayed healthcare-seeking behaviour significantly increased the risk of typhoid complications, while the rural residence appeared to be protective. These findings underscore the need for timely diagnosis, prompt treatment, and health education, especially in urban areas. Strengthening preventive strategies, including vaccination, remains essential to reducing typhoid-related complications and mortality.

## Introduction

Fever is among the most common symptoms reported by patients seeking healthcare, especially in poor-resource countries, and may occur either in isolation or alongside other symptoms [1]. Several febrile illnesses, including typhoid fever (hereafter referred to interchangeably as typhoid), are life-threatening conditions [2,3]. Typhoid, caused by *Salmonella enterica* serotype Typhi (*S.* Typhi), is a systemic febrile disease specific to humans. It is transmitted through the ingestion of water or food contaminated by the faeces of transient or chronic carriers. Annually, typhoid causes an estimated 11.9 to 26.9 million cases and 128 to 216.5 thousand deaths globally [4,5]. The disease remains a major public health concern, particularly in low- and middle-income countries, including Asia and Africa, where 18 million cases are reported each year [6].

In Ghana, typhoid ranked among the top 10 outpatient illnesses from 2020 to 2022, with 389 cases per 100,000 person-years of observation among children under 15 years [7,8]. Clinically, Typhoid presents with diverse signs and symptoms that mimic other febrile and gastrointestinal disorders, making accurate diagnosis challenging and increasing the risk of complications. Typhoid complications, including typhoid intestinal perforation (TIP), gastrointestinal bleeding, and encephalopathy, affect up to 15% of patients, particularly those with prolonged illness lasting over two weeks, with higher prevalence reported in Africa [9,10].

Several factors contribute to the increased risk of typhoid complications, including extreme age, male sex, antimicrobial resistance, prior antibiotic use, and delays in seeking medical care. Delayed medical intervention can lead to late diagnosis and treatment, further exacerbating the risk of complications. Effective management of typhoid hinges on understanding these risk factors, accurate diagnosis, timely antimicrobial therapy, and prevention through vaccination [11]. Prompt diagnosis and treatment are crucial to averting complications and reducing mortality [12].

While global data on typhoid complications are available, including from Africa [13,14], evidence from Ghana remains scarce regarding prevalence, types, and associated risk factors. To the best of our knowledge, typhoid intestinal perforation is the most frequently documented complication, while others may be underreported [15–21]. This study addresses a critical knowledge gap providing novel data on typhoid complications in Ghana and offering evidence to inform public health interventions and clinical management strategies. Specifically, we aimed to assess the prevalence, types, and risk factors of complications among febrile patients, with a focus on typhoid fever in the Ashanti region of Ghana. We hypothesised that delayed treatment due to prolonged symptoms before seeking healthcare increases the risk of complications.

## Methods

### Study settings

The study involved a retrospective analysis of surveillance data under the Severe Typhoid in Africa (SETA) program established in Ghana between May 2016 and September 2019. The design and methodology of the SETA study are published elsewhere [22]. Briefly, the SETA program conducted healthcare facility-based surveillance across Burkina Faso, the Democratic Republic of Congo, Ethiopia, Madagascar, Nigeria, and Ghana actively screening for invasive salmonellosis and typhoid fever complications using a standardised protocol. In Ghana, SETA exclusively recruited patients residing in the Ashanti region from two healthcare facilities: Komfo Anokye Teaching Hospital (KATH) in Kumasi and Agogo Presbyterian Hospital (APH) in Asante Akyem Agogo (Fig 1). APH is the primary referral hospital located in the Agogo township in the Asante Akim North district with a population of 85,788. The APH with a bed capacity of 350 and 125, 918 outpatient department attendances per year, provides healthcare services for patients all over Ghana and beyond [23,24]. KATH, a tertiary medical centre and teaching hospital situated in the Kumasi metropolis with a population of 443,981 [23,25], serves as the regional hospital for the Ashanti region and also sees patients from 11 other regions in Ghana. With a bed capacity of 1,200, it registers 40,000 in-patients and attends to 380,000 out-patient cases annually (KATH, 2019). The study obtained clinical and laboratory data from consenting eligible patients in the in-patient and out-patient units of KATH and APH hospitals. Blood cultures were used to confirm typhoid infection. Eligibility criteria included patients presenting with current fever (body temperature ≥38°C tympanic/rectal or ≥37.5°C axillary) or a history of fever lasting ≥3 consecutive days within the 7 days preceding enrollment.

**Fig 1.**
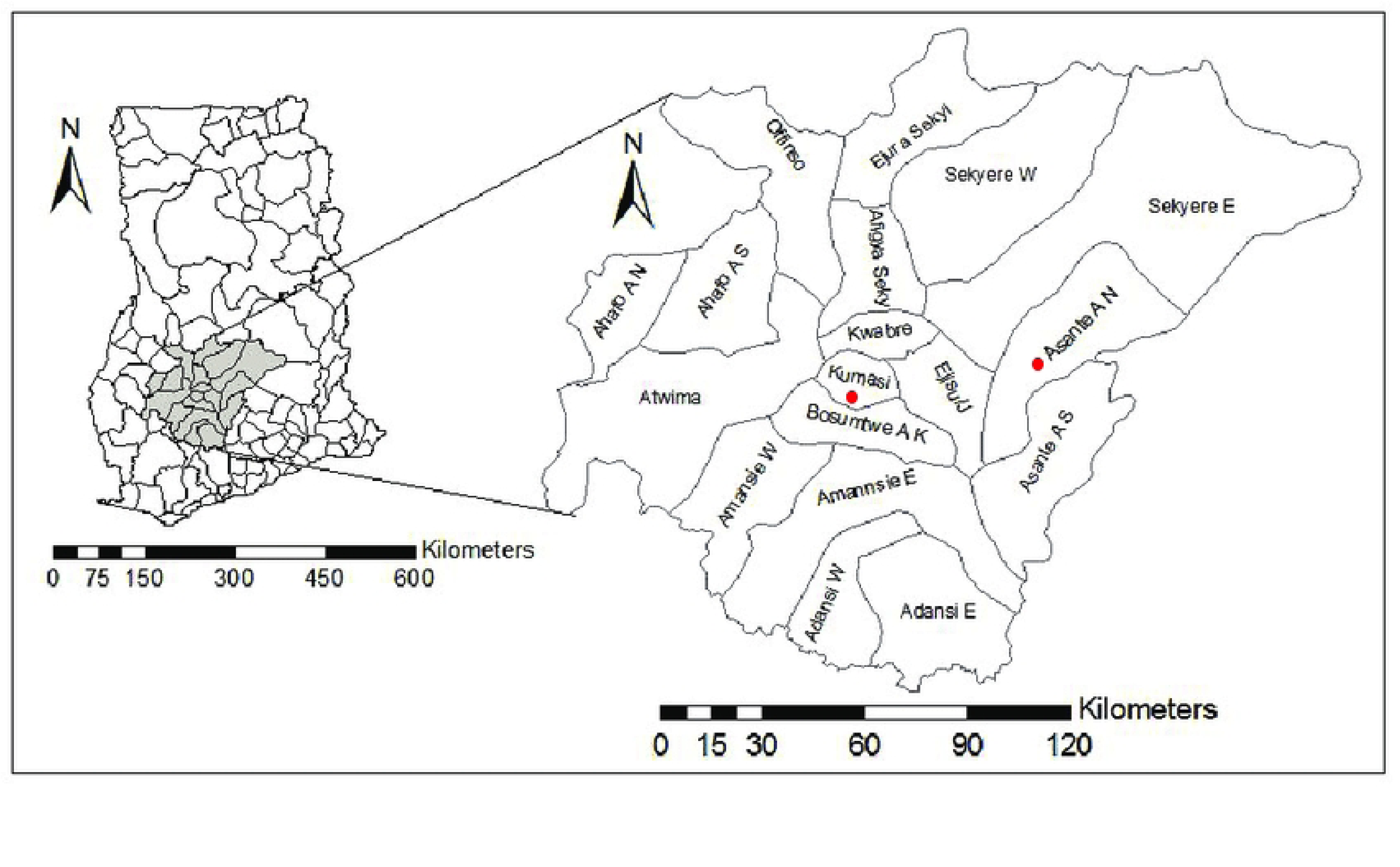
Map of Ashanti region, Ghana, indicating SETA study sites.

### Ethical approval

The SETA Ghana study protocol received ethical approval from the Committee on Human Research, Publication and Ethics (CHRPE) of the Kwame Nkrumah University of Science and Technology, School of Medical Sciences/Komfo Anokye Teaching Hospital. The initial approval was granted in April 2016 (CHPRE/AP/219/16) with renewals obtained throughout the data collection period (May 2016 to September 2019). Before enrollment, voluntary written informed consent was obtained from all participants, in the case of minors, from parents or legal guardians. For this study data were accessed in 20^th^ February 2023. The dataset was fully de-identified prior to access and the authors did not have access to any personally identifying information.

### Study variables

Using a standardised data abstraction form (S1 Appendix), data related to patients with positive blood culture growth were extracted from the SETA-Ghana database. The abstracted data contained a mix of continuous and categorical variables, covering demographic, clinical, and laboratory aspects for both inpatients and outpatients. The variables measured in days included: the duration of hospitalisation, duration of current fever, duration of past fever, duration of other symptoms before hospital visit (from the date fever started to the date of first healthcare assessment), and duration of illness before diagnosis (from the date symptoms started to the date blood culture ended). Based on clinical relevance to typhoid complication risk factors and a review of the literature, the following continuous data were categorised: an elevated white cell count (defined as >15×10^3^/uL), a low white cell count (defined as <4×10^3^/L), a low platelet count (defined as <140×10^3^/uL), low haemoglobin (Hb) (Hb <10g/dL in children ≤5 years or Hb <11.0g/dL in older children and adults).

In this study, we defined complications as the occurrence of one or more severe systemic conditions, including gastrointestinal bleeding (visible blood/melena or positive blood stool test), intestinal perforation (suspected or confirmed at surgery), encephalopathy (delirium, obtundation, stupor, or coma), meningitis (suggestive symptoms, CSF culture examination), myocarditis (abnormal cardiac rhythm and ventricular failure with an associated abnormality on the ECG or ultrasound evidence of a pericardial effusion),hepatitis (visible jaundice, hepatomegaly, elevated serum glutamic-oxaloacetic transaminase (SGOT >400 IU/L) and/or elevated serum glutamic-pyruvic transaminase (SGPT >400 IU/L)), cholecystitis (right upper quadrant pain and tenderness without evidence of hepatitis, enlarged/thickened wall gall bladder), pneumonia (respiratory symptoms, abnormal chest X-ray infiltrates), severe anemia (hematocrit <20%), focal infection (abscess drainage culture), renal impairment (creatinine ≥175μmol/L), a clinical and/ laboratory diagnosis of sepsis, gastritis, other liver diseases, appendicitis, central nervous system infection (CNS), requiring blood transfusion, or resulting in death.

### Microbiological methods

Under sterile conditions, 5-8 mL of blood from children and 10-14 mL from adults were drawn into BACTEC bottles (Becton-Dickenson, USA) at enrollment. The BACTEC bottles were incubated for up to 5 days in BACTEC 9050 automated incubator. Vials which flagged positive cultures underwent Gram staining and were plated on blood, chocolate or MacConkey agar. Gram-negative isolates were identified using API-20E and API-NH (Biomérieux, France), while Gram-positive *Staphylococcus aureus* was confirmed via coagulase testing. Antimicrobial susceptibility testing followed a modified Bauer-Kirby disc diffusion method, with inhibition zones interpreted according to the 2015 Clinical and Laboratory Standards Institute (CLSI) guidelines [26]. The antimicrobials tested, included: ampicillin (10µg), chloramphenicol (30µg), cotrimoxazole (1.25/23.75µg), ciprofloxacin (5µg), cefoxitin (30µg), ceftazidime (30µg), ceftriaxone/cefotaxime (30µg), cefuroxime (30µg), nalidixic acid (30µg) and tetracycline (30µg). Isolates were stored in microbank beads (Pro-labs Diagnostics Inc, UK) at −80°C and later sub-cultured on nutrient agar. Minimum inhibitory concentrations MICs for ciprofloxacin (0.008μg/mL to 4μg/mL) and nalidixic acid (0.5μg/mL to 512μg/mL) were determined using E-test strips (Biomérieux, France) per 2015 CLSI guidelines. *Escherichia coli* ATCC^®^ 25922 and *Staphylococcus aureus* ATCC^®^ 25923 served as control strains. An isolate was classified as multi-drug resistant (MDR) if it was resistant to all three antibiotics: ampicillin, chloramphenicol and cotrimoxazole, also known as first-line antibiotics against *S.* Typhi infection. In addition, it was defined as other multiply resistant if it showed resistance to at least two antibiotics. Resistance was determined using CLSI breakpoints, with intermediate and resistant categories classified as resistant.

### Selection criteria for study participants

Between 2016 and 2019, 2,196 febrile patients were recruited, and blood samples collected from 2174. Among these, 244 had positive blood culture growths. In total, 238 participants were included in the statistical analysis, with 6 excluded due to missing enrollment information. Seventy-two typhoid patients were identified. Of the participants whose cultures showed bacterial growth (244), 131 experienced at least one complicated health condition, with 28 cases resulting from those with typhoid fever, Fig 2.

**Fig 2.**
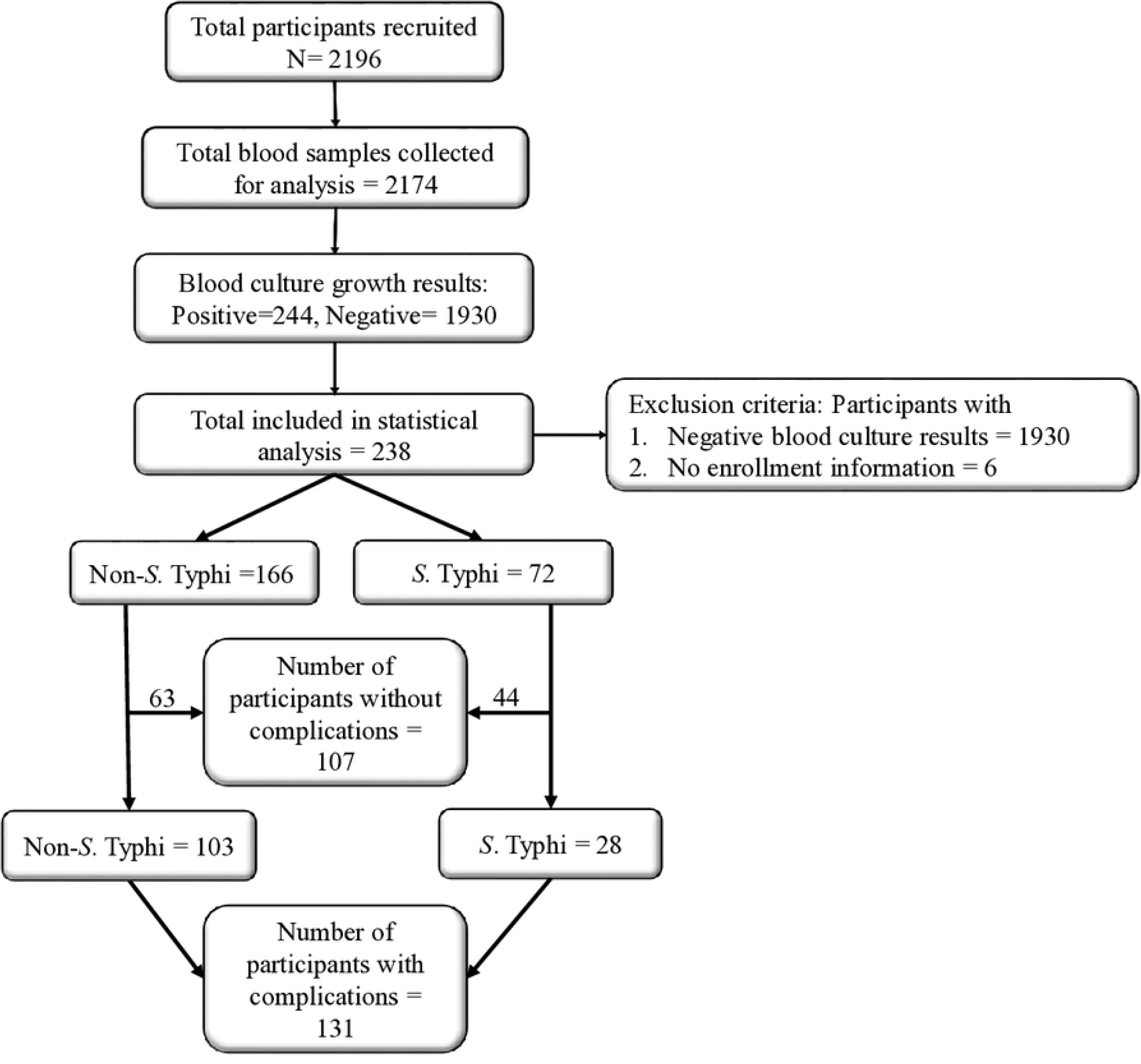
Selection of study participants.

### Statistical Analysis

All variables considered in this study were subjected to statistical data reduction techniques to avoid over-parameterisation, which can undermine inference quality due to too many explanatory variables [27]. We employed factor analysis with Promax rotation for continuous data reduction. For categorical data, we applied a chi-square test of independence, with a statistical significance of p≤0.05, using the patient’s complication status as the dependent variable. The reduced data were then analysed and presented in tables and figures stratified by causative agents (non-*S.* Typhi and *S.* Typhi) and across specific age groups (≤ 1 year, 2-5 years, 6–15 years, and ≥ 16 years). Continuous data were summarised with the median and interquartile range and compared using the Mann-Whitney U-test. Proportions were assessed using the Chi-squared test or Fisher’s exact test as appropriate. For logistic regression analysis, continuous variables including fever days, other symptoms days and days ill before diagnosis were categorised based on whether they fell below the median of their respective combined distributions. Univariate logistic regression identified risk factors and reported them as odds ratios (OR) with 95% confidence intervals (95% CI). Variables significant at p<0.10 in the typhoid univariate analyses were included in a multivariate model. Independent factors associated with typhoid complications were determined using a backward stepwise approach, with variables in the final model significant at p<0.05. All analyses were conducted using STATA version 14 (StataCorp, Texas, USA).

### Identification of key risk factors

A total of 58 variables, comprising 20 continuous and 38 categorical data, were included in the data reduction analysis using factor analysis and the chi-square test of independence.

### Component factor analysis for continuous variables

Summary statistics of the variables used in the factor analysis indicated that none followed a normal distribution (p< 0.05) (Table 1 in S2 Appendix). A preliminary inspection of the Spearman correlation matrix identified 21 significant correlations among the variables (Table 2 in S2 Appendix). Although all correlation coefficients remain <1, collinearity was detected between the variables “current fever” and “history of fever,” with a Rho=1.000. Consequently, “current fever” was excluded from further analysis. Subsequent Kaiser-Meyer-Olkin (KMO) tests were conducted to evaluate the data suitability for factor analysis [28,29]. The results initially fell short of the acceptable range with KMO^1^=0.44, which is below the adequacy criterion of 0.5, both overall and individually. The dataset eventually achieves an acceptable overall KMO value of 0.61 at KMO^2^. However, the variable “creatinine” fails to meet the adequacy criterion with KMO = 0.38. Finally, an overall KMO value of 0.65 was achieved at KMO^3^, indicating the data appropriateness for factor analysis with 14 factors despite initial shortcomings (Table 3 in S2 Appendix). Moreover, Bartlett’s test showed that the correlations, when taken collectively, are significant at the p<0.001. Table 4 in S2 Appendix lists the eigenvalues for the 14 possible factors, and based on the Kaiser rule of retaining factors with eigenvalues >1.0, four factors were retained. These four factors explain 98% of the total variance of the 14 factors, demonstrating that they sufficiently capture the underlying data structure. Therefore, these four factors were retained for further analysis. We then examined the factor pattern in Table 5 in S2 Appendix for significant factor loadings. The Promax rotation technique was applied to improve interpretation since the unrotated factor matrix did not show a clear factor loading pattern. In the rotated factor solution, each variable had a significant loading (>0.60) on only one factor, except for Respiratory rate (RESRT), Platelets (PLT), and SGPT, which had no significant loadings on any of the four factors considered. Consequently, these variables with insignificant loadings (RESRT, PLT, and SGPT) were removed from the analysis, leaving 11 variables for further consideration. The factor analysis was then re-conducted with the remaining 11 variables. Applying the Kaiser rule and inspecting the scree plot led to the retention of three factors (Fig. 1 in S2 Appendix), which accounted for 93% of the variance. This high level of explained variance indicates that these three factors provide a robust and simplified model for identifying risk factors for typhoid complications in Ghana, and are sufficient for capturing the essential information from the original variables.

**Table 1.** Distribution of study characteristics among 238 participants stratified by age groups.

| Variable | 0-1 year<br>n=60 | 2-5 years<br>n=54 | 6-15 years<br>n=86 | $\geq 16$ years<br>n=38 | All ages<br>(n=238) | P-value<sup>a</sup> | P-value<sup>b</sup> |
| --- | --- | --- | --- | --- | --- | --- | --- |
| *Hospital site<sup>c</sup>* |  |  |  |  |  | **0.001** | **<0.001** |
| APH (Rural) | 16/60(12.1) | 28/54(21.2) | 52/86(39.4) | 36/38(27.3) | 132/238(55.5) |  |  |
| KATH (Urban) | 44/60(41.5) | 26/54(24.5) | 34/86(32.1) | 2/38(1.9) | 106/238(44.5) |  |  |
| *Treatment delay* |  |  |  |  |  |  |  |
| Days fever prior to facility visit<sup>d</sup> | 3(2-7) | 4(2-6) | 5(3-7) | 3(2-5) | 4(2-7) | **0.002** | 0.102 |
| Days of symptoms | 4(3-7) | 4(2-6) | 6(3-7) | 3(2-5) | 4(2-7) | **0.003** | 0.062 |
| before hospital visit<sup>d</sup> |  |  |  |  |  |  |  |
| Days ill before diagnosis<sup>d</sup> | 7(5-11) | 8(5-10) | 9(6-11) | 9(6-11) | 8(6-11) | **0.007** | 0.571 |
| *Vital signs* |  |  |  |  |  |  |  |
| Weight<sup>d</sup> | 8.3(6-10) | 13.1(11.2-15.1) | 24.8(20-29.4) | 71(67-85) | 15(10-23) | **<0.001** | **0.001** |
| BP systolic (<90 or >120 mmHg)<sup>e</sup> | 7/13(53.9) | 21/25(84.0) | 27/47(57.5) | 10/25(40.0) | 65/110(59.1) | 0.123 | **0.027** |
| BP diastolic (<45 or >100 mmHg)<sup>e</sup> | 6/13(46.2) | 14/25(56.0) | 26/47(55.3) | 9/25(36.0) | 55/110(50.0) | 0.759 | 0.111 |
| Comorbidities | 20/60(33.3) | 11/54(20.4) | 16/86(18.6) | 5/38(13.2) | 52/238(21.9) | 0.075 | 0.157 |
| *Prior use of medication* |  |  |  |  |  |  |  |
| Antibiotics | 25/60(41.7) | 14/54(25.9) | 21/86(24.4) | 2/38(5.3) | 62/238(26.1) | 0.164 | **0.001** |
| Other drug<sup>f</sup> | 11/60(18.3) | 8/54(14.8) | 17/86(19.8) | 6/38(15.8) | 42/238(17.7) | 0.971 | 0.743 |
| *Signs & symptoms* |  |  |  |  |  |  |  |
| Abdominal pain | 2/60(3.3) | 7/54(13.0) | 34/86(39.5) | 7/38(18.4) | 50/238(21.0) | **<0.001** | 0.669 |
| Diarrhoea | 14/60(23.3) | 10/54(18.5) | 15/86(17.4) | 5/38(13.2) | 44/238(18.5) | 0.865 | 0.356 |
| Shortness of breath | 49/60(81.7) | 46/54(85.2) | 78/86(90.7) | 36/38(94.7) | 209/238(87.8) | **0.046** | 0.186 |
| Visible blood in the stool | 1/60(1.7) | 2/54(3.7) | 1/86(1.2) | 1/38 (2.6) | 5/238 (2.1) | >0.999 | 0.584 |
| Confusion | 0 | 1/54(1.9) | 3/86(3.5) | 1/38(2.63) | 5/238(2.1) | 0.348 | 0.584 |
| Hepatomegaly | 11/60(18.3) | 13/54(24.1) | 23/86(26.7) | 0 | 47/238(19.8) | 0.220 | **<0.001** |
| Splenomegaly | 6/60(10.0) | 9/54(16.7) | 9/86(10.5) | 0 | 24/238(10.1) | 0.794 | **0.018** |
| Altered consciousness | 2/60(3.3) | 1/54(1.9) | 2/86(2.3) | 2/38(5.3) | 7/238(2.9) | >0.999 | 0.310 |
| Prostration/exhaustion | 9/59(15.25) | 8/52(15.38) | 11/86(12.79) | 3/36(8.33) | 31/233(13.3) | 0.540 | 0.432 |
| *Hematological profile* |  |  |  |  |  |  |  |
| Low Hb<sup>#</sup> | 37/59(62.7) | 35/52(67.3) | 55/80(68.8) | 15/34(44.1) | 142/225(63.1) | 0.801 | **0.013** |
| WBC count |  |  |  |  |  |  |  |
| <4×10<sup>3</sup>/uL | 1/59(1.7) | 3/52(5.8) | 17/80(21.3) | 7/34(20.6) | 28/225(12.4) | **<0.001** | **0.033** |
| >15×10<sup>3</sup>/uL | 23/59(39.0) | 13/52(25.0) | 15/80(18.8) | 3/34(8.8) | 54/225(24.0) |  |  |
| *Bacterial isolate* |  |  |  |  |  | **<0.001** | 0.178 |
| Non-*S. Typhi* | 58/60(96.7) | 40/54(74.1) | 38/86(44.2) | 30/38(78.9) | 166/238(69.8) |  |  |
| *S. Typhi* | 2/60(3.3) | 14/54(25.9) | 48/86(55.8) | 8/38(21.1) | 72/238(30.3) |  |  |

Note: <sup>a</sup> Comparison of participants <5 years with 5-15 years old. <sup>b</sup> of <16 and ≥16 years old using Mann Whitney U, Chi-square or Fisher's exact tests where appropriate; P-value set at 5% significant level;
<sup>c</sup> Denominators of age categories are totals of each site; AAA-Asanti Akim Agogo; <sup>d</sup> Median (IQR); <sup>e</sup> Vital signs categorized based on the participant's age; <sup>f</sup> Other drugs include NSAID, analgesics and antipyretics, antimalarials, herbals etc.

**Table 2.** Antimicrobial susceptibility profile of isolates among 238 participants stratified by age groups.

| **Antibiotic resistance profile** | **0-1 year (N=60)** | **2-5 years (N=54)** | **6-15 years (N=86)** | **≥16 years (N=38)** | **All ages (N=238)** | **P-value<sup>a</sup>** | **P-value<sup>b</sup>** |
| --- | --- | --- | --- | --- | --- | --- | --- |
| Ampicillin | 13/22(59.1) | 16/28(57.1) | 20/59(33.9) | 15/24(62.5) | 64/133(48.1) | **0.026** | 0.119 |
| Chloramphenicol | 10/23(43.5) | 14/26(53.9) | 22/60(36.7) | 13/32(40.6) | 59/141(41.8) | 0.192 | 0.874 |
| Cotrimoxazole | 9/23(39.1) | 12/24(50.0) | 19/55(34.6) | 16/28(57.1) | 56/130(43.1) | 0.337 | 0.090 |
| Amoxicillin/Clavulanic acid | 9/19(47.4) | 3/20(15.0) | 12/54(22.2) | 9/22(40.9) | 33/115(28.7) | 0.171 | 0.159 |
| Cefoxitin | 0 | 0 | 1/4(25.0) | 1/2(50.0) | 2/8(25.0) | >0.999 | 0.464 |
| Ceftazidime | 6/22(27.3) | 1/24(4.2) | 3/55(5.5) | 3/55(13.6) | 13/123(10.6) | **0.043** | 0.701 |
| Ceftriaxone/Cefotaxime | 9/22 (40.9) | 3/26 (11.5) | 6/56(10.7) | 4/26 (15.4) | 22/130 (16.9) | **0.038** | >0.999 |
| Cefuroxime | 3/7(42.9) | 1/2(50.0) | 3/5(60.0) | 3/6(50.0) | 10/20(50.0) | 0.592 | >0.999 |
| Ciprofloxacin | 4/24(16.7) | 5/27(18.5) | 14/57(24.6) | 9/23(39.1) | 32/131(24.4) | 0.512 | 0.071 |
| Nalidixic acid | 0 | 2/19(10.5) | 4/46(8.7) | 1/11(9.1) | 7/83(8.4) | >0.999 | >0.999 |
| Penicillin | 3/7(42.9) | 1/2(50.0) | 3/5(60.0) | 3/6(50.0) | 10/20(50.0) | 0.592 | >0.999 |
| Tetracycline | 4/19(21.1) | 9/25(36.0) | 12/53(22.6) | 18/26(69.2) | 43/123(35.0) | 0.825 | **<0.001** |
| MDR | 4/21(19.1) | 9/23(39.1) | 14/53(26.4) | 10/21(47.6) | 37/118(31.4) | 0.889 | 0.079 |
| Other multiple | 14/24(58.3) | 15/28(53.6) | 27/61(44.3) | 21/32(65.6) | 77/145(53.1) | 0.155 | 0.108 |
| resistant ($\geq 2$ antibiotics) | | | | | | | |
| Fully susceptible organism | 4/24(16.7) | 7/28(25.0) | 23/61(37.7) | 6/32(18.8) | 40/145(27.6) | **0.020** | 0.205 |

Note: <sup>a</sup> Comparison of participants <5 years with 5-15 years old. <sup>b</sup> of <16 and $\geq 16$ years old using Mann Whitney U, Chi-square or Fisher's exact tests where appropriate; P-value set at 5% significant level; Other multiple resistant was when isolate is resistance to $\geq 2$ antibiotics.

**Table 3.** Complications among 238 study participants categorised by age group and bacterial isolates.

| Specific Complications | ≤15 years old |  | >15 years old |  | All ages |  |  |
| --- | --- | --- | --- | --- | --- | --- | --- |
|  | Non-*S. Typhi* (N=136) | *S. Typhi* (N=64) | Non-*S. Typhi* (N=30) | *S. Typhi* (N=8) | Non-*S. Typhi* (N=166) | *S. Typhi* (N=72) | Overall prevalence |
| Prevalence of complication | 93 (68.4) | 25 (39.1) | 10 (33.3) | 3 (37.5) | 103 (62.1) | 28 (38.9) | 131 (55.0) |
| Sepsis | 39 (28.7) | 11 (17.2) | 4 (13.3) | 2 (25.0) | 43 (25.9) | 13 (18.1) | 56 (23.5) |
| Gastritis | 0 | 0 | 0 | 1 (12.5) | 0 | 1 (1.4) | 1 (0.4) |
| Appendicitis | 2 (1.5) | 2 (3.1) | 0 | 0 | 2 (1.2) | 2 (2.8) | 4 (1.7) |
| Central nervous system infection | 16 (11.8) | 1 (1.6) | 2 (6.7) | 0 | 18 (10.8) | 1 (1.4) | 19 (8.0) |
| Cholecystitis | 1 (0.7) | 0 | 1 (3.3) | 0 | 2 (1.2) | 0 | 2 (0.8) |
| Gastrointestinal bleeding | 3 (2.2) | 6 (9.4) | 0 | 0 | 3 (1.8) | 6 (8.3) | 9 (3.8) |
| Intestinal perforation | 1 (0.7) | 4 (6.2) | 1 (3.3) | 0 | 2 (1.2) | 4 (5.6) | 6 (2.5) |
| Encephalopathy | 4 (2.9) | 0 | 1 (3.3) | 0 | 5 (3.0) | 0 | 5 (2.1) |
| Meningitis | 3 (2.2) | 0 | 0 | 0 | 3 (1.8) | 0 | 3 (1.3) |
| Myocarditis | 0 | 0 | 1 (3.3) | 0 | 1 (0.6) | 0 | 1 (0.4) |
| Hepatitis | 1 (0.7) | 1 (1.6) | 1 (3.3) | 0 | 2 (1.2) | 1 (1.4) | 3 (1.3) |
| Other liver diseases | 12 (8.8) | 2 (3.1) | 1 (3.3) | 0 | 13 (7.8) | 2 (2.8) | 15 (6.3) |
| Pneumonia | 14 (10.3) | 2 (3.1) | 2 (6.7) | 1 (12.5) | 16 (9.6) | 3 (4.2) | 19 (8.0) |
| Severe Anemia (Hct < 20%) | 27.94 | 4 (6.3) | 4 (13.3) | 0 | 42 (25.3) | 4 (5.6) | 46 (19.3) |
| Focal infection | 10 (7.4) | 1 (1.6) | 0 | 1 (12.5) | 10 (6.0) | 2 (2.8) | 12 (5.0) |
| Renal impairment | 4 (2.9) | 1 (1.6) | 5 (16.7) | 0 | 9 (5.4) | 1 (1.4) | 10 (4.2) |
| Blood transfusion | 20 (14.7) | 2 (3.1) | 3 (10.0) | 0 | 23 (13.9) | 2 (2.8) | 25 (10.5) |
| Death | 3 (2.2) | 1 (1.6) | 1 (3.3) | 0 | 4 (2.4) | 1 (1.4) | 5 (2.1) |

**Table 4.**
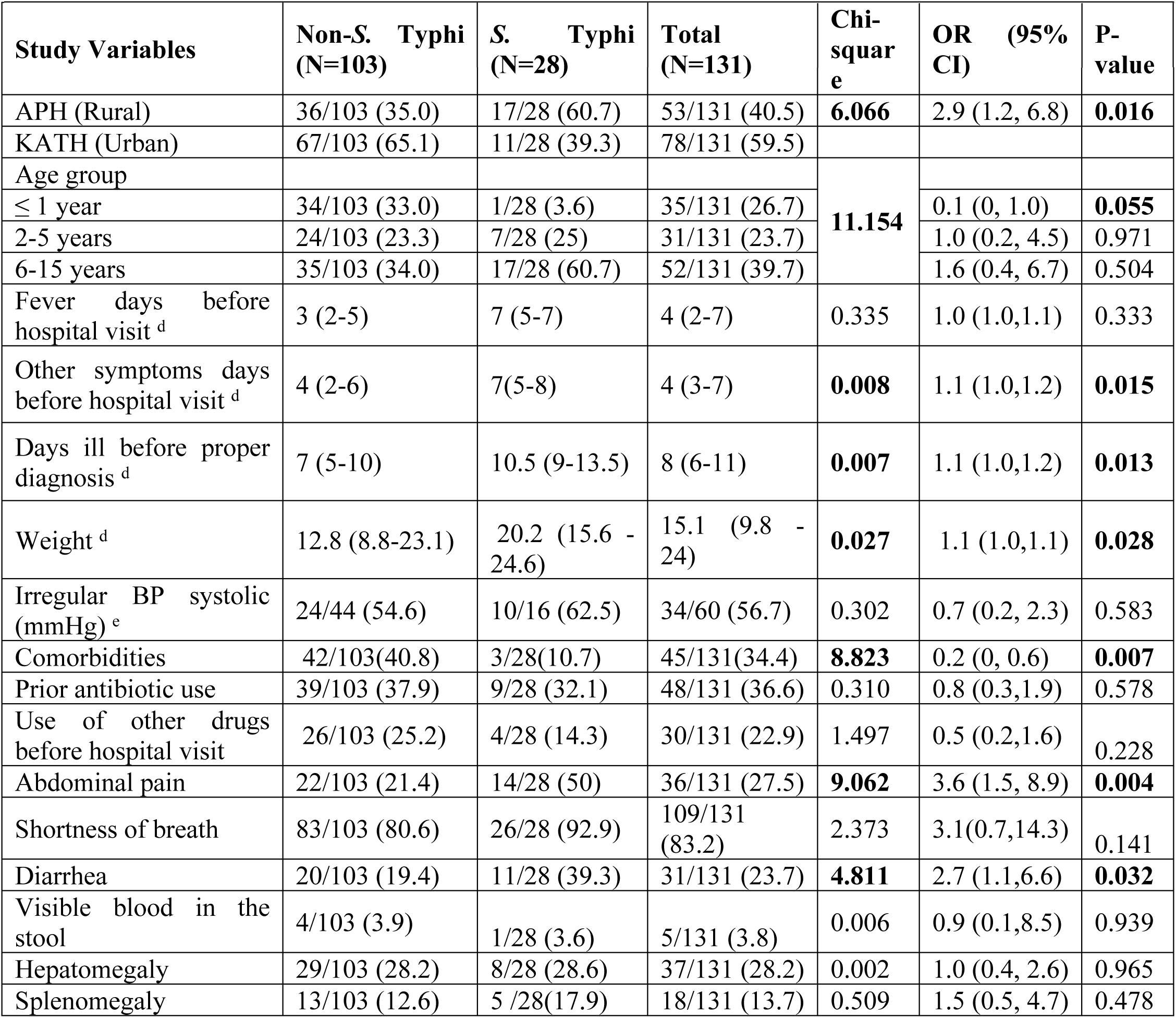
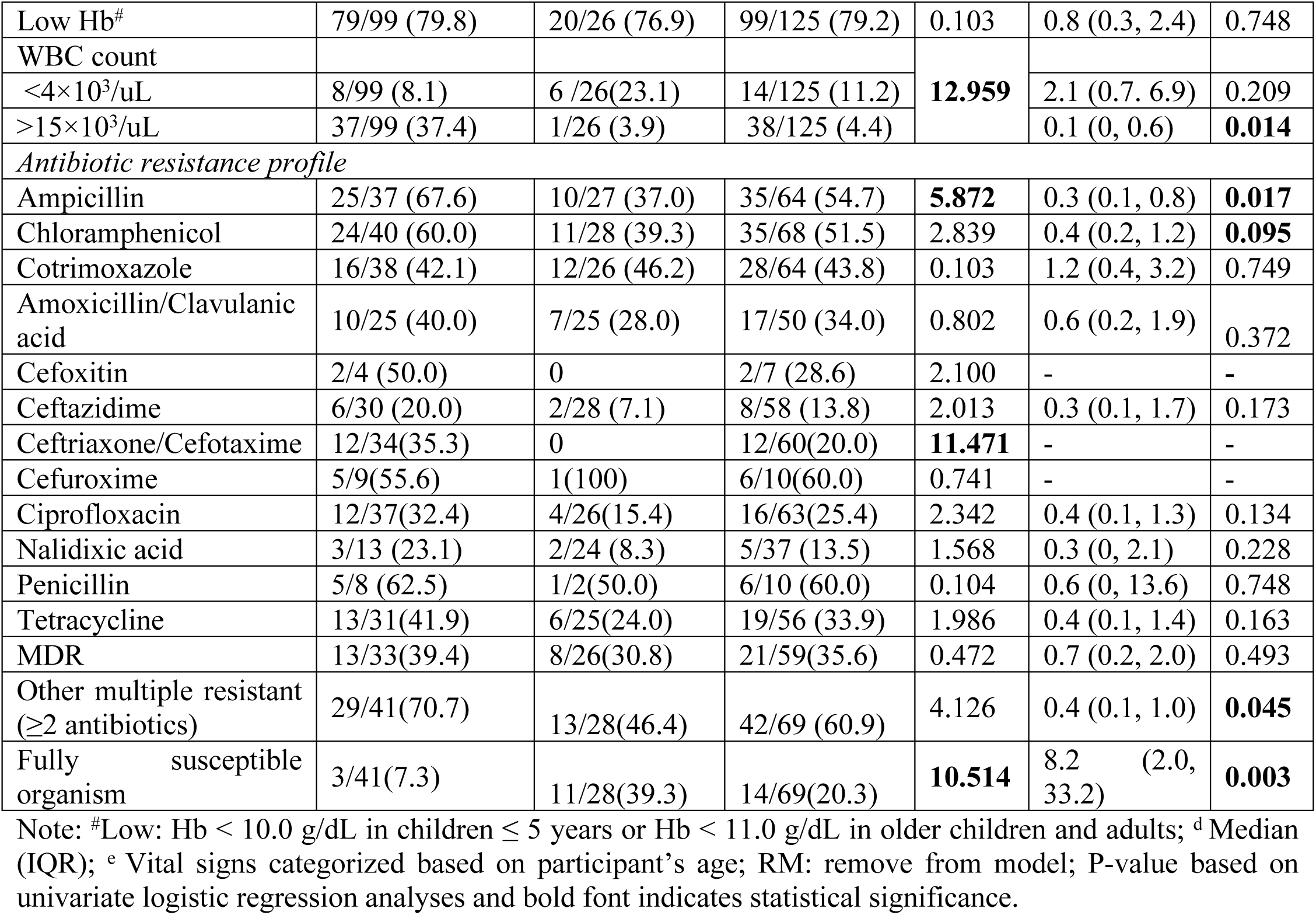
Characteristics of febrile patients with complications grouped by non*-S.* Typhi and *S.* Typhi.

**Table 5.** Risk factors for *S*. Typhi complications. Logistic regression models.

| Study Variables | No complication (N=44) | Complications (N=28) | OR (95% CI) | P-value | AOR (95% CI) | P-value |
| --- | --- | --- | --- | --- | --- | --- |
| APH (Rural) | 43/44 (97.7) | 17/28 (60.7) | 0.04 (0, 0.3) | **0.002** | 0.1 (0, 0.7) | **0.024** |
| KATH (Urban) | 1/44 (2.3) | 11/28 (39.3) |  |  |  |  |
| *Treatment delay* |  |  |  |  |  |  |
| Fever days before hospital visit <sup>a</sup> |  |  |  |  |  |  |
| $\geq 6$ days | 18/37 (48.7) | 19/27 (70.4) | 2.5 (1.0, 7.1) | 0.086 | RM | |
| Days of other symptoms before hospital visit <sup>a</sup> |  |  |  |  |  |  |
| $\geq 6$ days | 17/37 (45.9) | 20/27 (74.1) | 3.4 (1.1, 9.9) | **0.027** | 8.6 (1.2, 64.7) | **0.035** |
| Days ill before proper diagnosis <sup>a</sup> |  |  |  |  |  |  |
| $\geq 10$ days | 19/44 (43.2) | 19/28 (67.9) | 2.8 (1.0, 7.5) | **0.044** | 0.3 (0, 2.2) | 0.225 |
| Weight <sup>d</sup> | 23.2 (19.3, 32.7) | 20.2 (15.6, 24.6) | 0.9 (0.9, 1.0) | 0.054 | RM |  |
| *Prior use of medication* |  |  |  |  |  |  |
| Antibiotics | 4/44 (9.1) | 9/28 (32.1) | 4.7 (1.3, 17.4) | **0.019** | 1.5 (0.1, 15.8) | 0.740 |
| Other drugs | 1/44 (2.3) | 4/28 (14.3) | 7.2 (0.8, 67.8) | 0.086 | RM |  |
| *Sign & Symptoms* |  |  |  |  |  |  |
| Abdominal pain | 11/44 (25.0) | 14/28 (50.0) | 3.0 (1.1, 8.2) | **0.033** | 0.6 (0.1, 2.6) | 0.453 |
| Shortness of breath | 42/44 (95.4) | 26/28 (92.9) | 0.6 (0.1, 4.7) | 0.642 |  |  |
| Diarrhoea | 7/44 (15.9) | 11/28 (39.3) | 3.4 (1.1, 10.4) | **0.030** | 2.7 (0.6, 12.8) | 0.214 |
| Hepatomegaly | 4/44 (9.1) | 8/28 (28.6) | 4.0 (1.1, 14.9) | **0.039** | 1.6 (0.2, 13.2) | 0.679 |
| Splenomegaly | 4/44 (9.1) | 5/28 (17.9) | 2.2 (0.3, 8.9) | 0.281 |  |  |
| *Antibiotic resistance profile* |  |  |  |  |  |  |
| Ampicillin | 7/43 (16.3) | 10/27 (37.0) | 3.0 (1.0, 9.3) | 0.054 | RM |  |
| Chloramphenicol | 8/42 (19.1) | 11/28 (39.3) | 2.8 (0.9, 8.0) | 0.067 | RM |  |
| Cotrimoxazole | 8/39 (20.5) | 12/28 (46.2) | 3.3 (1.1, 9.9) | **0.032** | RM |  |
| Amoxicillin/Clavulanic acid | 5/43 (11.6) | 7/25 (28.0) | 3.0 (0.8, 10.6) | 0.096 | RM |  |
| Ciprofloxacin | 9/42 (21.4) | 4/26 (15.4) | 0.7 (0.2, 2.4) | 0.540 |  |  |
| Nalidixic acid | 1/41 (2.4) | 2/24 (8.3) | 3.6 (0.3, 42.4) | 0.303 |  |  |
| Tetracycline | 7/41 (17.1) | 6/25 (24.0) | 1.5 (0.4, 5.2) | 0.494 |  |  |
| MDR | 7/39 (18.0) | 8/26 (30.8) | 2.0 (0.6, 6.5) | 0.234 |  |  |
| Other multiple resistance ($\geq 2$) | 8/43 (18.6) | 13/28 (46.4) | 3.7 (1.3, 11.0) | **0.014** | 3.5 (0.6, 19.4) | 0.149 |
| *Hematological profile* |  |  |  |  |  |  |
| Low Hb # | 17/42 (40.5) | 20/26 (76.9) | 4.9 (1.6, 14.7) | **0.005** | 2.6 (0.5, 14.1) | 0.257 |
| WBC count |  |  |  |  |  |  |
| $<4 \times 10^3/\mu\text{L}$ | 9/42 (21.4) | 6/26 (23.1) | 1.1 (0.3, 3.5) | 0.889 | | |
| $>15 \times 10^3/\mu\text{L}$ | 2/42 (4.8) | 1/26 (3.9) | 0.8 (0.1, 9.6) | 0.872 | | |

#Low: Hb < 10.0 g/dL in children $\leq 5$ years or Hb < 11.0 g/dL in older children and adults; <sup>a</sup> Categorized based on the Median; <sup>b</sup> Vital signs categorized based on participant's age; OR: crude odds ratio; AOR: Adjusted odds ratio; 95% CI: 95% confidence interval; RM: removed from model.

Next, we named each factor based on the variables with significant loadings from Table 6 in S2 Appendix (Promax rotated factor pattern). Factor 1, which loads highly onto AGE, systolic BP (BPSYS), diastolic (BPDIA), and weight (WGHT) was named “Risk of hypertension.” Factor 2, with significant loadings on white blood cells (WBC), hemoglobin (Hb), and hematocrit (HCT), was termed “hematological profile.” Finally, Factor 3, based on significant loadings from duration of other symptoms (SYMPD), duration of fever before the hospital visit (FEVERD), and duration of illness before proper diagnosis (TRTD), was named “Treatment delay.” We used these 10 variables as surrogates in further analysis, however, they retained their original names in case any were found to be significantly associated with typhoid fever complications.

### Chi-square test of independence for categorical data

Excluding antimicrobials, we applied Pearson’s chi-square test to 38 categorical variables to identify key variables associated with the occurrence of complications (grouped as Yes/No) (as detailed in Table 7 in S2 Appendix). Variables with a p≤0.10 were considered statistically significant. These included “hospital site,” “comorbidities,” “pathogenicity (*S.* Typhi),” “drug exposures before hospital visit (antibiotics and other drugs),” and “signs and symptoms (such as abdominal pain, shortness of breath, diarrhoea, visible blood in stool and prostration or exhaustion)”.

## Results

### Pathogen Distribution

This study identified various pathogens and contaminants among participants with complications, showing a significant association with all ages (p=0.001). Pathogens included *S.* Typhi (21.4%, 28/131), *Salmonellae* Typhimurium (0.8%, 1/131), other non-typhoidal *Salmonellae* (9.2%, 12/131), *Staphylococcus aureus* (4.6%, 6/131), *E. coli* (3.8%, 5/131), *Klebsiella* spp. (3.1%, 4/131) and *Streptococcus pneumoniae* (2.3%, 3/131). Contaminants were Coagulase-Negative *Staphylococci* (CNS) (33.6%, 44/131), *Micrococcu*s spp. (3.8%, 5/131), *Bacillus* sp. (3.1%, 4/131), and *Viridans streptococci* (3.1%, 4/131), etc. (Fig 3).

**Fig 3.**
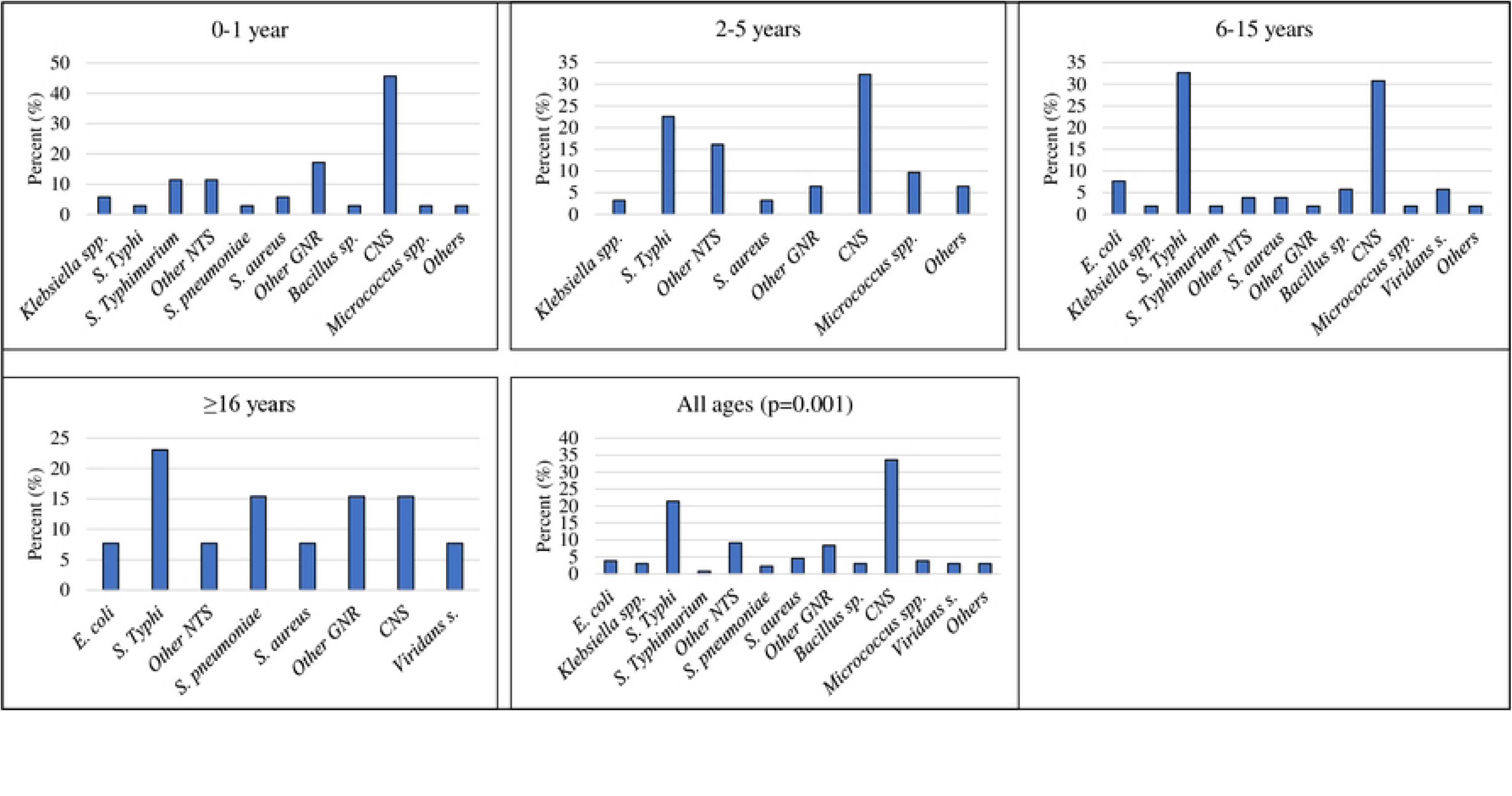
The distribution of pathogens and contaminants among participants with complications. NTS: Non-Typhoidal *Salmonellae* GNR: Gram-Negative Rods Others: Other Contaminants, including *Weeksella virosa, Listeria* spp., R*alstonia picketti, Raoultella ornithinolytica*, ect.)

### Description of the reduced variables

Further analyses of the reduced dataset encompassed 28 variables (10 continuous and 18 categorical) involving 238 participants, predominantly 84% (200/238) children (<16 years) and 16% (38/238) adults (≥16 years). Table 1 outlines the distribution of study characteristics stratified by age groups. Hospital site distribution showed notable disparities, with a higher representation of 6-15-year-olds (39.4%, 52/132) in the rural (APH) and 0-1-year-olds (41.51%, 44/106) in urban (KATH) settings. Children aged 6-15 years exhibited prolonged febrile illness durations before seeking medical attention, with median delays of 5 days for fever symptoms, 6 days for other symptoms, and 9 days before diagnosis. BPSYS irregularities were significantly more prevalent in children compared to adults (p=0.027), notably affecting 84% (21/25) of those aged 6-15 years. The median weight varied significantly across age groups: 8.3 kg for 0-1 years, 24.8kg for 6-15 years, and 71kg for older patients. Comorbidities were also evidently higher among children, with 33.3% (20/60) prevalence in the 0-1 age group. Antibiotic use before hospital visits was more significant in children (<16 years) than in older patients (p=0.001). Among 6-15-year-olds, abdominal pain was prevalent in about 40% (34/86), while shortness of breath was even more common, affecting almost 91% (78/86). Hepatomegaly and splenomegaly occurred exclusively in children <16 years, with prevalences of 26.7% (23/86) in those aged 6-15 years and 16.7% (9/54) in those aged 2-6 years, respectively. Pathogen isolate distribution showed a significant predominance of *S.* Typhi among children <16 years, particularly in the 6-15-year age group (55.8%, 48/86).

### Antibiotic susceptibility profile of isolates

Table 2 outlines the antimicrobial susceptibility profile of organisms isolated across different age groups. Approximately, 28% (40/145) of all isolates were fully susceptible to the tested antimicrobials. In children aged 6-15, 62.5% (15/24) of the isolates exhibited significant resistance to ampicillin (p=0.026), whereas resistance to chloramphenicol or cotrimoxazole was not statistically significant in any age group. Moreover, the prevalence of multidrug resistance (MDR) was highest in the 6-15 age group (26.4%, 14/53), followed by the >16 years age group (47.6%, 10/21). Among isolates from children aged 0-1 year, 27.3% (6/22) were significantly resistant to ceftazidime (p=0.043), while 40.9% (9/22) showed resistance to ceftriaxone/cefotaxime (p=0.038). In contrast, more than 69% (18/26) of isolates from patients older than 16 years demonstrated a strong association with tetracycline resistance (p<0.001). Of the 145 isolates, 77 (53.1%) were resistant to two or more antibiotics, with the highest burden observed in children aged 6-15 years 27/61(44.3).

### Prevalence of complications

Table 3 summarises disease complications across age groups of bacterial isolates. Overall, 55% (131/238) of patients experienced complications, with 49.6% (118/238) occurring in children aged ≤15 years and 5.5% (13/238) in those >15 years. *S.* Typhi and non-*S.* Typhi caused 38.9% (28/72) and 62.1% (103/166) of these complications, respectively. Notably, sepsis, not previously listed as a typhoid complication by Parry, et al. [14], was observed in 18.1% (13/72) of typhoid cases, affecting 17.2% (11/64) of children ≤15 years and 25% (2/8) of those >15 years. In non-*S*. Typhi cases, sepsis occurred in 29% (39/136) of children ≤15 years and 13% (4/30) in those >15 years. Other complications in *S.* Typhi cases included gastrointestinal bleeding (8.3%, 6/72), intestinal perforation (5.6%, 4/72), hepatitis (1.4%, 1/72), and pneumonia (4.2%, 3/72). Non-*S.* Typhi patients had these complications at 1.8% (3/166), 1.2% (2/166), 1.2% (2/166), and 9.6% (16/166). The majority of *S.* Typhi-related gastrointestinal bleeding (9.4%, 6/64) and intestinal perforation (6.2%, 4/64) occurred in children ≤15 years. Cholecystitis, meningitis, myocarditis, and encephalopathy were exclusively observed in non-*S.* Typhi cases with prevalence of 1% (2/166), 1.8% (3/166), 0.6% (1/166), and 3% (5/166), respectively. Severe anaemia was more common in non-*S.* Typhi cases (25%, 42/166) compared to *S.* Typhi cases (5.6%, 4/72). Similarly, renal impairment was more prevalent in non-*S*. Typhi 5.4% (9/166) than in *S.* Typhi 1.4% (1/72). (Blood transfusions were required in 2.8% (2/72) of *S.* Typhi cases and 13.9% (23/166) of non-*S.* Typhi cases, predominantly in children ≤15 years. The overall mortality rate across all age groups was 2.1% (5/238), with 1.4% (1/72) of these deaths occurring in a child with an *S.* Typhi infection. However, the cause of death specifically related to typhoid was not determined in this study.

### Comparison of complications in *S.* Typhi and non-*S.* Typhi febrile patients

Distinct characteristics of *S.* Typhi versus (vs) non-*S.* Typhi in febrile patients with complications are detailed in Table 4. *S.* Typhi complications were more common in rural areas (60.7%, 17/28), while non-*S.* Typhi complications were more frequent in urban areas (65.1%, 67/103). Both were notably represented in the 6-15-year age group (61% for *S.* Typhi, 34% for non-*S*. Typhi). *S.* Typhi patients experienced longer delays before treatment, with a median illness duration of 10.5 days before diagnosis. They also had a longer duration of fever (7 vs. 3 days) and other symptoms (7 vs. 4 days) before hospital visits compared to patients with non-*S.* Typhi complications. Non-*S.* Typhi isolates from patients with complications detected resistance to ampicillin (67.6%, 25/37), chloramphenicol (60%, 24/40), and cotrimoxazole (42.1%, 16/38), while *S.* Typhi isolates exhibited resistance to ampicillin (37%, 10/27), chloramphenicol (39.3%, 11/28), and cotrimoxazole (46.2%, 12/26). Cefoxitin (50%, 2/4) and ceftriaxone (35.3%, 12/34) resistance occurred exclusively in non-*S*. Typhi isolates, with one *S*. Typhi isolate resistant to cefuroxime. Ciprofloxacin and nalidixic acid resistance were higher in non-*S*. Typhi (32.4% and 23.1%) than in *S.* Typhi (15.4% and 8.3%). Tetracycline resistance was detected in 42% (13/31) of non-*S.* Typhi and 24% (6/25) of *S.* Typhi isolates. While MDR was not statistically significant (p = 0.493), other multiple resistance was significantly higher (p = 0.045) in non-*S.* Typhi (70.7%, 29/41) than in *S*. Typhi (46.4%, 13/28). *S.* Typhi isolates showed greater overall susceptibility (39.3%, 11/28) compared to non-*S.* Typhi (7.3%, 3/41) isolates.

### Risk factors associated with typhoid fever complications

Table 5 shows the logistic regression analyses and the distribution of typhoid cases with and without complications. Of the 72 typhoid fever patients, 28 had complications, and 44 did not. Complications were significantly more frequent in patients from rural areas (60.7%, 17/28) than from urban areas (39.3%, 11/28). However, patients from rural areas were less likely to develop complications than urban patients (OR 0.04, 95% CI 0-0.3, p=0.002). Patients with fever for ≥6 days had 2.5 times higher odds of developing complications compared to those with shorter fever durations (<6 days) (OR 2.5, 95% CI 1.0-7.1, p=0.086). Similarly, delays in seeking treatment for other symptoms were associated with the risk of typhoid complications, with patients experiencing longer durations (≥6 days) having 3.4 times higher odds of complications (OR 3.4, 95% CI 1.1-9.9, p=0.027). Furthermore, patients with complications were more likely to have taken antibiotics before hospital visits (OR 4.7, 95% CI 1.3-17.4, p=0.019), have hepatomegaly (OR 4.0, 95% CI 1.1-14.9, p=0.039), and present with diarrhoea (OR 3.4, 95% CI 1.1-10.4, p=0.030) and abdominal pain (OR 3.0, 95% CI 1.1-8.2, p=0.033). Low haemoglobin levels were also significantly associated with complications (OR 4.9, 95% CI 1.6-14.7, p=0.005). Antibiotic resistance factors, including ampicillin (OR 3.0, 95% CI 1.0-9.3, p=0.054), chloramphenicol (OR 2.8, 95% CI 0.9-8.0, p=0.067), nalidixic acid (OR 3.6, 95% CI 0.3-42.4, p=0.303) and multidrug resistance (OR 2.0, 95% CI 0.6-6.5, p=0.234) increased the odds of typhoid complications. However, these factors were not statistically significant at the 95% CI, except for cotrimoxazole (OR 3.3, 95% CI 1.1–9.9, p = 0.039) and other multiple resistance (resistant to ≥2 antibiotics) (OR 3.7, 95% CI 1.3–11.0, p = 0.014). Additionally, patients with low Hb were about 5 times more likely to develop complications than those with normal Hb (OR 4.9 95% CI 1.6-14.7). After adjusting for potential factors in the multivariate model, typhoid complications were independently associated with longer delays in treatment (symptoms duration before hospital visit) (AOR 8.6, 95% CI 1.2-64.7, p=0.032) and the location of healthcare seeking (AOR 0.1, 95% CI 0-0.7, p=0.024). Although not statistically significant, the following factors indicated an increased odds of typhoid fever complications: diarrhoea (OR 2.7 95% CI 0.6-12.8, p=0.214), low Hb (OR 2.6 95% CI 0.5-14.1, p=0.257) and isolate resistant to two or more antibiotics (multiple resistant) (OR 3.5 95% CI 0.6-19.4, p=0.149)

## Discussion

### Burden of Febrile illness complications

In this study, we examined 238 febrile patients with positive blood cultures from rural and urban hospitals in the Ashanti region of Ghana. We observed a general disease complication rate of 55% (131/238), which is significantly higher than the 28% to 30% range reported in previous studies [30–32]. This higher prevalence observed in our study could likely be attributed to differences in research methodologies, study populations, and environmental contexts. Our study employed a more stringent approach to identifying and classifying pathogenic bacteria, with a particular emphasis on enhancing the detection of invasive *Salmonella* infections. In contrast, previous studies focused on a broader range of febrile illnesses or specific organ-related complications. Furthermore, our research encompassed patients of all age groups from resource-limited settings in both rural and urban areas, whereas earlier studies primarily concentrated on urban populations and were often limited to children or adults [30,31].These variations in methodology, population, and environmental factors likely account for the observed differences in the febrile complication rates.

### Occurrence of typhoid fever-related complications

Among the 238 febrile patients studied, 72 (30.3%) had typhoid fever. Of the 131 patients with complications, 28 (21.4%) were attributed to *S.* Typhi. Notably, 38.9% (28/72) of typhoid fever patients who were hospitalized or attending hospitals experienced complications. This rate exceeds previous reports, such as the 10-15% prevalence noted by Crump, et al. [9], 27% by Cruz Espinoza, et al. [33], and 26% by Marchello, Birkhold and Crump [10]. The most common typhoid complication was sepsis, affecting 18.1% (13/72) of patients, followed by gastrointestinal bleeding at 8.3% (6/72), intestinal perforation at 5.6% (4/72), and severe anaemia also at 5.6% (4/72). Sepsis being the predominant complication aligns with studies by Mahle and Levine [34] and Parry, et al. [35], who suggests that young children often exhibit non-specific symptoms of syndromic sepsis in the advanced stages of typhoid fever, making early diagnosis challenging. The prevalence of gastrointestinal bleeding among *S.* Typhi patients reported in our study also aligns with a study by Crump, et al. [9], which reported occurrences in up to 10% of patients. However, our observed prevalence of intestinal perforation exceeded the previously reported range of 1% to 3% by Parry, et al. [35] and Crump, et al. [9]. Severe anaemia was notably prevalent among patients with complications, which may be attributed to factors such as intestinal bleeding, nutritional deficiencies, and hemolysis linked to prevalent malaria in West Africa. We also observed an increased risk of complications associated with symptoms such as abdominal pain, diarrhoea, and hepatomegaly, which are recognised indicators of typhoid infection [9]. Mortality occurred in a patient with gastrointestinal bleeding from *S.* Typhi infection at a rate of 1.4% (1/72), closely corresponding to the 1% case fatality rate estimated by Crump, Luby and Mintz [36] and Buckle, Walker and Black [37]. This rate also aligns with the estimates of 0.4% to 2.1% provided by Mogasale, et al. [4] and Marchello, Birkhold and Crump [10].

Our findings also underscore an independent association between delays in treatment and the occurrence of typhoid complications, supporting previous studies that demonstrate the detrimental effects of delayed antimicrobial therapy on complication rates [38,39]. Studies reported that the timing, dosage, and duration of antimicrobial therapy are critical for preventing complications in typhoid fever [10,33]. Early initiation of treatment at the appropriate dose and for the correct duration is essential. In developing countries, delays in seeking treatment are often due to poor health-seeking behaviour [40] and the overlap with malaria, which can present similarly to typhoid fever. Specifically, this study observed that over 74% of patients with a median disease duration of ≥6 days experienced complications, showing about 9 times (AOR 8.6, 95% CI 1.2-64.7), the risk of complications compared to those who sought treatment within a median duration of <6 days. Similar research in Africa indicates that delays ranging from 6 to 11 days before initiating treatment are associated with a higher risk of complications [10,33,41]. Such delays not only increase the likelihood of severe outcomes, including higher mortality rates, extended hospital stays, and complications like organ failure or abscesses, but also exacerbate the progression of infections, inflate healthcare costs, and contribute to growing antimicrobial resistance [35,42].

We also discovered an association between typhoid complications and both prior antibiotic use (OR 4.7, 95% CI 1.3-17.4, p=0.019), and resistance to ≥2 antibiotics (multiply resistance) (OR 3.7, 95% CI 1.3-11.0, p=0.014) using univariate models. However, these associations did not reach statistical significance in the multivariate analysis, resulting in adjusted odds ratios of 1.5 (95% CI 0.1-15.8, p=0.740) for prior antibiotic use and 3.5 (95% CI 0.6-19.4, p=0.149) for multiple resistance. Despite the lack of statistical significance in the multivariate model, the persistently increased risk may be attributed to the indiscriminate use of antibiotics, which is often a result of poor health-seeking behaviours in many developing countries, including Ghana. Previous research highlights that unnecessary antimicrobial exposure can foster drug resistance, increasing the risk of MDR and extensively drug-resistant (XDR) infections [43,44]. Although this study found no direct association between MDR phenotype and typhoid complications, patients with MDR had a twofold higher risk of complications compared to those without resistance (OR 2.0, 95% CI 0.6-6.5, p=0.234). In addition, a significant association was observed between cotrimoxazole, a first-line antibiotic, and typhoid complications (OR 3.3, 95% CI 1.1-9.9, p=0.032). Resistance of *S.* Typhi isolates to first-line antibiotics and MDR has been widely documented in several African countries, including Ghana [20]. Typhoid conjugate vaccines (TCVs) have proven effective in preventing typhoid fever and could be instrumental in limiting *S.* Typhi transmission while decreasing antibiotic use [45]. Moreover, TCVs could help prevent complications, thereby mitigating the severity of typhoid.

Another notable factor in this study was the relationship between hospital locations and both the prevalence and severity of typhoid infection. When analysed by age groups, patients aged 2 years and older experienced a higher disease burden in rural areas than in urban settings. This reinforces our overall SETA study findings across six African countries, which highlight a greater *S.* Typhi burden in rural areas compared to urban settings [20]. Furthermore, more patients seeking healthcare in rural areas presented with typhoid complications than those in urban areas. However, patients treated at the rural hospital had a significantly lower risk of typhoid complications than those in urban settings (AOR 0.1, 95% CI 0–0.7, p= 0.024). Reversing the reference to rural in this analysis revealed that patients in urban hospitals have higher adjusted odds of developing typhoid complications than those in rural areas (AOR 19.0, 95% CI 1.5-246.5, p=0.024). Typically, urban areas have more healthcare facilities and better access to medical resources, which should facilitate earlier detection and treatment of typhoid, potentially reducing complications. However, the increased risk of complications observed in the urban hospital may stem from differences in healthcare-seeking behaviour or referral patterns, where more severe cases are directed to urban centres.

### Study limitation and strength

The findings of this study should be interpreted in light of certain limitations. First, the data were drawn exclusively from patients who were hospitalised or received care at two referral hospitals in the Ashanti region. This could potentially introduce selection bias, as individuals who did not access these facilities, due to financial barriers, geographic inaccessibility or preference for traditional medicine, were not captured. Consequently, the results may not be fully representative of the wider population, particularly in rural and underserved areas. In addition, the relatively small sample size may limit the generalizability of the findings, warranting cautious interpretation. For the future, studies with larger sample size that extends beyond the two hospitals and employs a recruitment strategy integrating both hospital-/clinic-based and community-focused approaches are recommended to validate these findings and uncover additional risk factors for typhoid-related complications.

Despite these limitations, the study has several strengths. It leverages laboratory-confirmed diagnoses from a well-defined surveillance program (SETA), enhancing diagnostic accuracy. The use of rigorous statistical methods also strengthens the reliability of the associations found between clinical complications and specific risk factors. Moreover, this study provides novel data on typhoid complications in Ghana, addressing a critical knowledge gap and offering evidence to inform public health interventions and clinical management strategies.

## Conclusion

This study reveals a significant burden of disease complications among febrile patients, especially those with typhoid fever in Ghana. Treatment delays significantly increased the risk of typhoid complications, while the rural residence was independently associated with complications. In resource-limited settings, typhoid fever remains a critical cause of mortality. Given the similarity of typhoid fever symptoms with other febrile illnesses, it is appropriate for enhanced education on the infection with improved advocacy for early testing before treatment to tackle delays that will lead to a rise in related complications. Moreover, programmes should be targeted at urban communities to unravel the drivers of health-seeking behaviours concerning febrile conditions. Also, an enhanced focus on vaccines would be a strategic effort to prevent typhoid complications and reduce mortality.

## Data Availability

The data underlying this study cannot be shared publicly due to consortium-level data governance and restrictions outlined in the data sharing policy of the Severe Typhoid in Africa (SETA) surveillance program. However, qualified researchers who meet the criteria for access to confidential data may request access from the Principal Investigator in Ghana via email at.

## Acknowledgement

Special thanks to the EOD project team at Kwame Nkrumah University of Science in collaboration with the International Vaccine Institute for their support.

## Author contributions

All authors contributed equally to this manuscript. All authors contributed to the manuscript’s drafting, reviewing and editing for publication. All authors agreed and approved the final manuscript for publication.

## Supporting information

**S1 Appendix.** Data abstraction form

**S2 Appendix.** This file contains the output of the statistical data reduction techniques employed in this study

## References

1. Crump JA, Newton PN, Baird SJ, Lubell Y. Febrile Illness in Adolescents and Adults. In: Holmes KK BS, Bloom BR, Jha P, ed. Major Infectious Diseases. 3rd ed ed. Washington (DC): The International Bank for Reconstruction and Development / The World Bank; 2017:Chapter 14.

2. Havelaar AH, Kirk MD, Torgerson PR, Gibb HJ, Hald T, Lake RJ, et al. World Health Organization Global Estimates and Regional Comparisons of the Burden of Foodborne Disease in 2010. PLoS Med. 2015; 12(12). e1001923. DOI:10.1371/journal.pmed.1001923. PMC4668832 the World Health Organization advisory body-the Foodborne Disease Burden Epidemiology Reference Group-without remuneration. The authors declare no competing interests.

3. Jensenius M, Han PV, Schlagenhauf P, Schwartz E, Parola P, Castelli F, et al. Acute and potentially life-threatening tropical diseases in western travelers—a GeoSentinel multicenter study, 1996– 2011. Am J Trop Med Hyg. 2013; 88(2). 397.

4. Mogasale V, Maskery B, Ochiai RL, Lee JS, Mogasale VV, Ramani E, et al. Burden of typhoid fever in low-income and middle-income countries: a systematic, literature-based update with risk-factor adjustment. Lancet Glob Health. 2014; 2(10). e570–580. DOI:10.1016/S2214-109X(14)70301-8,

5. Stanaway JD, Reiner RC, Blacker BF, Goldberg EM, Khalil IA, Troeger CE, et al. The global burden of typhoid and paratyphoid fevers: a systematic analysis for the Global Burden of Disease Study 2017. Lancet Infect Dis. 2019; 19(4). 369–381.

6. Antillón M, Warren JL, Crawford FW, Weinberger DM, Kürüm E, Pak GD, et al. The burden of typhoid fever in low-and middle-income countries: a meta-regression approach. PLoS Negl Trop Dis. 2017; 11(2). e0005376.

7. Marks F, von Kalckreuth V, Aaby P, Adu-Sarkodie Y, El Tayeb MA, Ali M, et al. Incidence of invasive salmonella disease in sub-Saharan Africa: a multicentre population-based surveillance study. Lancet Glob Health. 2017; 5(3). e310–e323. DOI:10.1016/S2214-109X(17)30022-0. PMC5316558

8. MOH (GH). Health sector annual program of work. 2022 holistic assessment report; Ministry of Health; Republic of Ghana. 2023; pg.7. Available at: https://www.moh.gov.gh/wp-content/uploads/2024/03/2022-Holistic-Assessment-Report_FINAL3.pdf.

9. Crump JA, Sjolund-Karlsson M, Gordon MA, Parry CM. Epidemiology, Clinical Presentation, Laboratory Diagnosis, Antimicrobial Resistance, and Antimicrobial Management of Invasive Salmonella Infections. Clin Microbiol Rev. 2015; 28(4). 901–937. DOI:10.1128/CMR.00002-15. PMC4503790

10. Marchello CS, Birkhold M, Crump JA. Complications and mortality of typhoid fever: A global systematic review and meta-analysis. J Infect. 2020; 81(6). 902–910. DOI:10.1016/j.jinf.2020.10.030. PMC7754788

11. Parry CM, Wijedoru L, Arjyal A, Baker S. The utility of diagnostic tests for enteric fever in endemic locations. Expert Rev Anti Infect Ther. 2011; 9(6). 711–725. DOI:10.1586/eri.11.47,

12. Kumar A, Roberts D, Wood KE, Light B, Parrillo JE, Sharma S, et al. Duration of hypotension before initiation of effective antimicrobial therapy is the critical determinant of survival in human septic shock. Crit Care Med. 2006; 34(6). 1589–1596.

13. Birkhold M, Coulibaly Y, Coulibaly O, Dembélé P, Kim DS, Sow S, et al. Morbidity and mortality of typhoid intestinal perforation among children in sub-Saharan Africa 1995–2019: a scoping review. World journal of surgery. 2020; 44(9). 2892–2902.

14. Parry CM, Hien TT, Dougan G, White NJ, Farrar JJ. Typhoid fever. N Engl J Med. 2002; 347(22). 1770–1782. DOI:10.1056/NEJMra020201,

15. Abantanga FA, Wiafe-Addai BB. Postoperative complications after surgery for typhoid perforation in children in Ghana. Pediatr Surg Int. 1998; 14(1-2). 55–58. DOI:10.1007/s003830050435,

16. Archampong EQ. Operative treatment of typhoid perforation of the bowel. Br Med J. 1969; 3(5665). 273–276. DOI:10.1136/bmj.3.5665.273. PMC1984075

17. Clegg-Lamptey JN, Hodasi WM, Dakubo JC. Typhoid ileal perforation in Ghana: a five-year retrospective study. Trop Doct. 2007; 37(4). 231–233. DOI:10.1258/004947507782332784,

18. Damien P, Nabare C, Baiden F, Kwara E, Apanga S, Etego-Amengo S. How are surgical theatres in rural Africa utilized? A review of five years of services at a district hospital in Ghana. Trop Doct. 2011; 41(2). 91–95.

19. Oheneh-Yeboah M. Postoperative complications after surgery for Typoid Ileal performation in adults in Kumasi. West Afr J Med. 2007; 26(1). 32–36.

20. Marks F, Im J, Park SE, Pak GD, Jeon HJ, Wandji Nana LR, et al. Incidence of typhoid fever in Burkina Faso, Democratic Republic of the Congo, Ethiopia, Ghana, Madagascar, and Nigeria (the Severe Typhoid in Africa programme): a population-based study. Lancet Glob Health. 2024; 12(4). e599–e610. DOI:10.1016/S2214-109X(24)00007-X. PMC10951957

21. Birkhold M, Datta S, Pak GD, Im J, Ogundoyin OO, Olulana DI, et al. Characterization of typhoid intestinal perforation in Africa: results from the severe typhoid fever surveillance in Africa Program. Open Forum Infect Dis. 2023; 10(Supplement_1). S67–S73.

22. Park SE, Toy T, Cruz Espinoza LM, Panzner U, Mogeni OD, Im J, et al. The Severe Typhoid Fever in Africa Program: Study Design and Methodology to Assess Disease Severity, Host Immunity, and Carriage Associated With Invasive Salmonellosis. Clin Infect Dis. 2019; 69(Suppl 6). S422–S434. DOI:10.1093/cid/ciz715. PMC6821161

23. GSS. Ghana 2021 population and housing census; General report. Ghana Statistical Service; Population of regions and districts. 2021; Volume 3A p.53–54. Available at: https://statsghana.gov.gh/gssmain/fileUpload/pressrelease/2021%20PHC%20General%20Report%20Vol%203A_Population%20of%20Regions%20and%20Districts_181121.pdf.

24. Yeboah C. Agogo Presby Hospital holds annual performance review meeting. Joy Online, Health. 2024 May 2; [Cited 2024 July 25]. Available from: https://www.myjoyonline.com/agogo-presby-hospital-holds-annual-performance-review-meeting/.

25. KATH. Health Agency; Komfo Anokye Teaching Hospital. 2024 [Cited 2024 July 26]; Ministry of Health, Republic of Ghana, A Healthy Population for National Development. Available at: https://www.moh.gov.gh/komfo-anokye-teaching-hospital/.

26. CLSI. Clinical and Laboratory Standards Institute: M100-S25. Performance standards for antimicrobial susceptibility testing; twenty-fifth informational supplement. CLSI. 2015; 35(1-240.

27. Harrell FE. Regression modeling strategies. New York: Springer; 2015.

28. Hair JF, Anderson RE, Tatham RL, Black WC. Multivariate data analysis. 4th ed. Upper Saddle River, New Jersey: Prentice-Hall, Inc; 1995.

29. Tabachnick B, Fidell L. Using multivariate statistics. Boston, MA: Pearson Education Inc; 2007.

30. Salagre KD, Sahay RN, Pazare AR, Dubey A, Marathe KK. A Study of Clinical Profile of Patients presenting with Complications of Acute Febrile Illnesses During Monsoon. J Assoc Physicians India. 2017; 65(9). 37–42.

31. Afzal Z, Kallumadanda S, Wang F, Hemmige V, Musher D. Acute febrile illness and complications due to murine typhus, Texas, USA. Emerg Infect Dis. 2017; 23(8). 1268.

32. Condra CS, Parbhu B, Lorenz D, Herr SM. Charges and complications associated with the medical evaluation of febrile young infants. Pediatr Emerg Care. 2010; 26(3). 186–191. DOI:10.1097/PEC.0b013e3181d1e180,

33. Cruz Espinoza LM, McCreedy E, Holm M, Im J, Mogeni OD, Parajulee P, et al. Occurrence of typhoid fever complications and their relation to duration of illness preceding hospitalization: a systematic literature review and meta-analysis. Clin Infect Dis. 2019; 69(Supplement_6). S435–S448.

34. Mahle W, Levine M. Salmonella typhi infection in children younger than five years of age. Pediatr Infect Dis J. 1993; 12(8). 627–631. 10.1097/00006454-199308000-000018414773

35. Parry CM, Thompson C, Vinh H, Chinh NT, Phuong le T, Ho VA, et al. Risk factors for the development of severe typhoid fever in Vietnam. BMC Infect Dis. 2014; 14(73. DOI:10.1186/1471-2334-14-73. PMC3923984

36. Crump JA, Luby SP, Mintz ED. The global burden of typhoid fever. Bull World Health Organ. 2004; 82(5). 346–353. PMC 2622843. PMC 2622843

37. Buckle GC, Walker CL, Black RE. Typhoid fever and paratyphoid fever: Systematic review to estimate global morbidity and mortality for 2010. J Glob Health. 2012; 2(1). 010401. DOI:10.7189/jogh.02.010401. PMC3484760

38. Adegoke SA, Ige JT, Mohammed LO, Oluwasuyi Ige E. Predictors of intestinal perforation in children with typhoid fever. J Pediatr Infect Dis. 2011; 6(4). 247–251.

39. Bulage L, Masiira B, Ario AR, Matovu JKB, Nsubuga P, Kaharuza F, et al. Modifiable risk factors for typhoid intestinal perforations during a large outbreak of typhoid fever, Kampala Uganda, 2015. BMC Infect Dis. 2017; 17(1). 641. DOI:10.1186/s12879-017-2720-2. PMC5613338

40. Tinuade O, Iyabo RA, Durotoye O. Health-care-seeking behaviour for childhood illnesses in a resource-poor setting. J Paediatr Child Health. 2010; 46(5). 238–242. DOI:10.1111/j.1440-1754.2009.01677.x,

41. Hoffner RJ, Slaven E, Perez J, Magana RN, Henderson SO. Emergency department presentations of typhoid fever. J Emerg Med. 2000; 19(4). 317–321. DOI:10.1016/s0736-4679(00)00260-2,

42. Kaljee LM, Pach A, Garrett D, Bajracharya D, Karki K, Khan I. Social and Economic Burden Associated With Typhoid Fever in Kathmandu and Surrounding Areas: A Qualitative Study. J Infect Dis. 2018; 218(suppl_4). S243–S249. DOI:10.1093/infdis/jix122. PMC6226633

43. Kamal R, Ching C, Zaman MH, Sultan F, Abbas S, Khan E, et al. Identification of multiple variant extensively drug-resistant typhoid infections across Pakistan. The American Journal of Tropical Medicine and Hygiene. 2023; 108(2). 278–284.

44. Fida S, Mansoor H, Saif S, Iqbal J, Khan AQ. Clinical Perspectives of Multiple and Extensively Drug-Resistant Typhoid; result from a tertiary care hospital from Pakistan. The Journal of Infection in Developing Countries. 2021; 15(04). 530–537.

45. Patel PD, Patel P, Liang Y, Meiring JE, Misiri T, Mwakiseghile F, et al. Safety and Efficacy of a Typhoid Conjugate Vaccine in Malawian Children. N Engl J Med. 2021; 385(12). 1104–1115. DOI:10.1056/NEJMoa2035916. PMC8202713

